# Analysis of the outcome of patients with stage IV uterine serous carcinoma mimicking ovarian cancer

**DOI:** 10.1101/19002410

**Authors:** Murad Al-Aker, Karen Sanday, James Nicklin

## Abstract

**Objectives:** To identify clinicopathological factors that might influence survival in patients with stage IV uterine serous carcinoma, and to compare survival outcomes in patients with stage IV uterine serous carcinoma managed with neoadjuvant chemotherapy followed by interval cytoreduction (with or without adjuvant chemotherapy), primary cytoreductive surgery followed by adjuvant chemotherapy.

**Methods:** A retrospective cohort study of all patients with stage IV Uterine serous carcinoma treated between 2005 and 2015 within a regional cancer centre. Progression-free and overall survival rates were calculated using the Kaplan–Meier method.

**Results:** Of 50 women with stage IV uterine serous carcinoma who met inclusion criteria, 37 underwent primary cytoreductive surgery, nine received neoadjuvant chemotherapy with planned interval cytoreductive surgery and four received palliative care only. A pre-treatment diagnosis of stage IV uterine serous carcinoma was made for only 45.9% of the primary cytoreductive surgery group and 56.6% of the neoadjuvant chemotherapy group, with advanced ovarian cancer the most common preoperative misdiagnosis. Median follow up was 19 months. Median overall survival was 27 months for the primary cytoreductive surgery group, 20 months for the neoadjuvant chemotherapy group and two months for the palliative care group. Optimal cytoreduction was achieved in 67.6% of the primary cytoreductive surgery group and 87.5% of the neoadjuvant chemotherapy group who underwent interval cytoreduction. Optimal cytoreduction was associated with improvement in overall survival, compared with suboptimal cytoreduction (36 versus 15 months; P=0.16). Adjuvant chemotherapy was associated with significantly higher overall survival compared with no adjuvant chemotherapy (36 versus four months; P<0.05). Median overall survival was 16 months for those with pure uterine serous carcinoma (n=40), compared with 32 months for those with mixed histopathology (n=10).

**Conclusion:** Stage IV uterine serous carcinoma can mimic advanced ovarian cancer. It carries a poor prognosis, which is worse for pure uterine serous carcinoma than for mixed-type endometrial adenocarcinoma. Neoadjuvant chemotherapy followed by interval cytoreduction and adjuvant chemotherapy seems to be a safe option, with an increased rate of optimal cytoreduction and comparable overall survival, compared with primary cytoreductive surgery. Adjuvant chemotherapy significantly improves survival in all groups.

**Primary objective:** To analyse the clinicopathological factors that might influence the progression-free survival and overall survival in patients with stage IV uterine serous carcinoma treated at Queensland Centre for Gynecological cancer.

**Secondary objective:** To compare the survival outcomes of patients with stage IV uterine serous carcinoma treated with neoadjuvant chemotherapy and interval cytoreduction, with those treated with primary cytoreductive surgery followed by adjuvant chemotherapy and patients who received palliative care only.

**PRECIS:** Optimal cytoreduction and adjuvant chemotherapy improved survival in stage IV uterine serous carcinoma. Neoadjuvant chemotherapy was feasible and safe. Patients with microscopic disease have similar poor prognosis.

**HIGHLIGHTS:**

- Pure uterine serous carcinoma carries a worse prognosis compared to mixed uterine serous carcinoma
- Optimal cytoreduction and adjuvant chemotherapy improve survival in Stage IV uterine serous carcinoma
- Neoadjuvant chemotherapy is feasible and a safe option in the management of stage IV uterine serous carcinoma

## INTRODUCTION

Uterine serous carcinoma is a biologically distinct histological subtype of uterine cancer, resembling serous adenocarcinoma of the ovary.^1,2^ It represents an estimated 10% of all endometrial cancer,^3^ but accounts for up to 50% of endometrial cancer-specific deaths due to its tendency toward early metastasis^4-6^ and a pattern of metastatic spread similar to that of epithelial ovarian cancers.^7,8^

Surgery is the mainstay of treatment, and optimal cytoreduction has been shown to correlate with prolonged survival.^9-11^ Patients are usually offered adjuvant chemotherapy.^12-14^ The role of whole-abdomen radiotherapy is still controversial.^15-17^

Neoadjuvant chemotherapy has also been proposed for advanced stage IV uterine serous carcinoma informed by protocols and experience described for advanced ovarian cancer. Reported safety and effectiveness outcomes of neoadjuvant chemotherapy are comparable to those of primary cytoreductive surgery.^18-24^

This study analysed outcomes for women with stage IV uterine serous carcinoma treated at the Queensland Centre for Gynaecological cancer, Queensland, Australia.

## Methods

### Study design

We performed a retrospective cohort study in a large centralised gynaecological oncology service with a large catchment area. The local institutional review board approved the study and was conducted according to the declaration of Helsinki and ethical clinical practice guidelines.

### Study population

The study included women aged 18 years and over with stage IV uterine cancer and a histopathology report of serous histology or mixed histology with more than 10% other than serous component, who underwent surgical tumour cytoreduction, received neoadjuvant chemotherapy, or was referred to palliative care between 1 January2005 and 31 December2014. Eligible subjects were identified from a database maintained within the department of gynaecological oncology at Royal Brisbane and Women’s Hospital as a part of Queensland Centre for Gynaecological cancer central electronic database.

All histopathology were reviewed at the time of diagnosis by an expert gynaecological histopathologist, and each woman’s diagnosis and stage were verified at a multidisciplinary team meeting. Women with no verified histopathology or stage, or those who were not evaluated or treated in Queensland Centre for Gynaecological cancer, were excluded from this analysis.

Patients were offered neoadjuvant chemotherapy at the discretion of the treating physician, after reviewing of histopathology at the multidisciplinary team meeting.

### Data collected

Data were collected for baseline demographics, comorbidity (Charlson comorbidity index), Eastern Cooperative Oncology Group performance status (Zubrod scale)^25^, the treatment offered (surgical cytoreduction, chemotherapy or palliative therapy) and histopathology.

Treatment outcomes included surgical cytoreduction status assessed as optimal (defined as no single area of disease with a diameter greater than 1cm at the end of the operation) or suboptimal (more than 1cm residual disease), response (assessed by imaging, CA125 levels), overall survival, and progression-free survival (assessed according to radiologic, pathologic or biochemical evidence of recurrent disease). Postoperative complications were collected, where reported.

### Statistical analysis

Kruskal-Wallis and Fisher exact tests were used to compare demographics and surgical outcomes. Overall survival and progression-free survival were estimated using the Kaplan-Meier method. Subgroups according to Histopathological diagnosis and treatment modality were compared by log-rank analysis, with significance defined as P=0.05.

## RESULTS

### Study population

Among 3793 patients treated for endometrial cancer, uterine serous carcinoma was diagnosed in 153 patients (4.0%) and stage IV uterine cancer in 131 patients (3.5%). Fifty patients (1.3%) met inclusion criteria, which were 32.7% of all patients diagnosed with uterine serous carcinoma and 38.2% of all with stage IV uterine cancer. The median follow up was 19 months for the entire cohort.

### Baseline characteristics

The median age for the cohort was 71years, and the median age for the palliative treatment group was higher at 77.8years, but it was not statistically significant. The percentage of women older than 80years was 16.2% in the primary cytoreductive surgery group and 33.3% in the neoadjuvant chemotherapy group. Charlson comorbidity index was greater than 3 in 77.8% of patients who received neoadjuvant chemotherapy, compared with 43.2% of those who underwent primary surgery (p=0.001). All patients who were transferred to palliative care had a Charlson comorbidity index of 3 or higher. Zubrod performance status score of 2 or greater was recorded for 22(59.5%) of patients who underwent primary surgery, compared with six patients (67%) of those who received neoadjuvant chemotherapy and 75% of the palliative care group. Median CA125 was 110 IU/L for the primary cytoreductive surgery group, 250 IU/L for the neoadjuvant chemotherapy group and 217.5 IU/L for the palliative care group. (Table1)

**Table 1:**
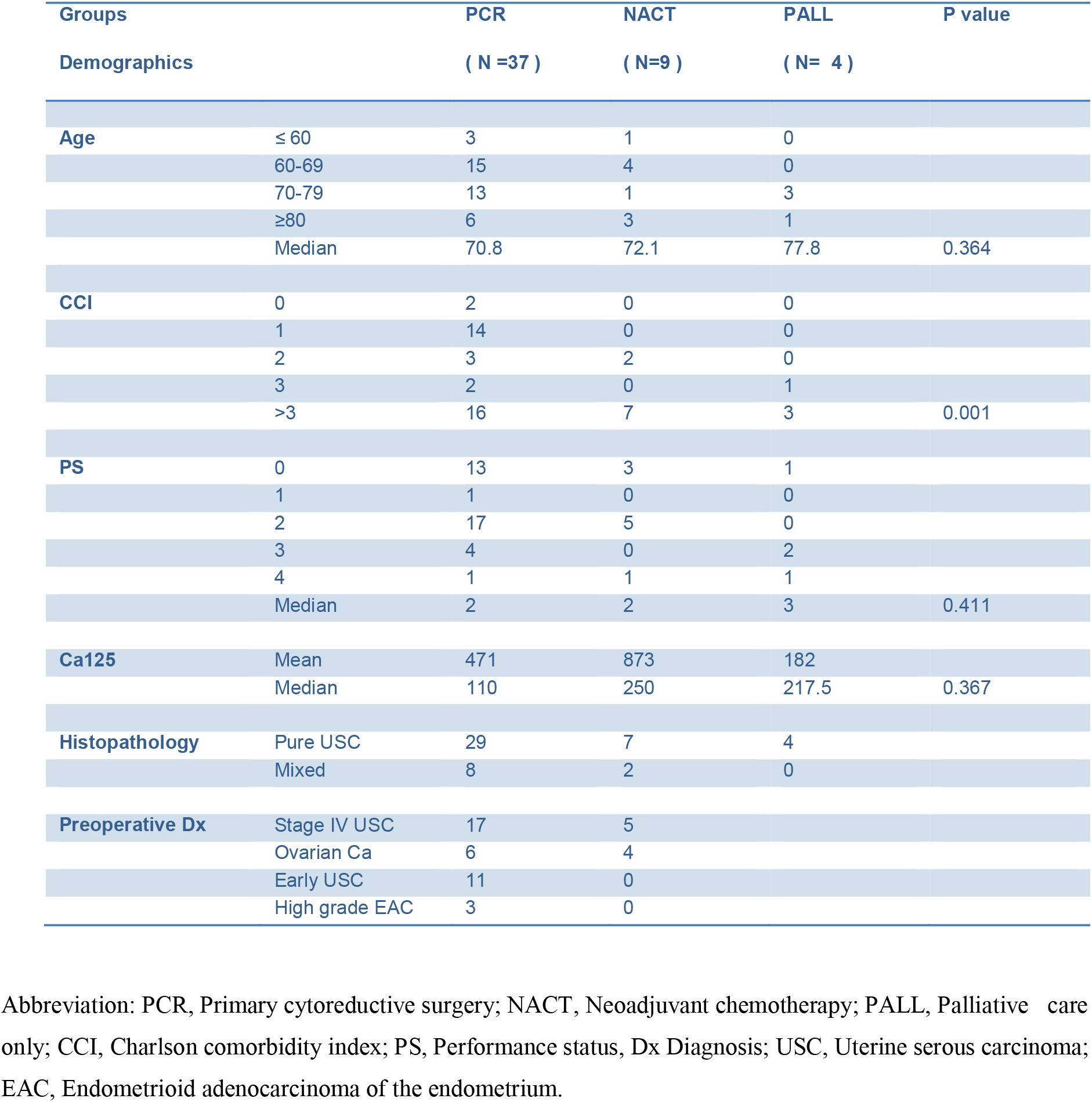
Stage IV Uterine serous carcinoma, Demographics, Ca125, preoperative diagnosis and histopathology.

### Treatment modalities

Thirty-seven patients underwent primary cytoreductive surgery, of whom 29 (78.4%) received adjuvant chemotherapy, and four (10.8%) received both adjuvant chemotherapy and radiotherapy. Of those who did not receive adjuvant treatments, one declined chemotherapy, five were assessed as too unwell to undergo chemotherapy and were transferred to palliative care, and two progressed within a short interval and did not receive treatment. Nine patients received neoadjuvant chemotherapy, of whom eight underwent interval cytoreductive surgery and received further chemotherapy. Four patients did not receive any disease-modifying treatment and were offered only palliative interventions.

### Histopathology

There were 40 cases of pure uterine serous carcinoma and 10 cases of mixed histopathology with >20% serous components. Of those with mixed histopathology, six patients (60%) had mixed endometrial adenocarcinoma and uterine serous carcinoma, two (20%) had mixed clear cell carcinoma and uterine serous carcinoma, and two (20%) had mixed undifferentiated carcinoma and uterine serous carcinoma. (Table1)

### Pre-surgical diagnosis

Advanced abdominal malignancy was diagnosed preoperatively in 26 patients who underwent primary cytoreductive surgery (70.3%). The preoperative stated intention was cytoreduction of stage IV uterine serous carcinoma in 17 patients (45.9%) and cytoreduction of advanced ovarian cancer in six patients (16.2%). Of those who received neoadjuvant chemotherapy, the stated intention of cytoreductive surgery was cytoreduction of advanced ovarian cancer in four patients (44.4%). In all these cases, uterine serous carcinoma was the final diagnosis at MDT, based on the histopathology.

Early uterine serous carcinoma was suspected, based on preoperative imaging, in 11 (29.7%) of the patients who underwent primary cytoreductive surgery. Of these women, six (16.2%) were found intraoperatively to have gross peritoneal/omental disease involvement, and the remaining five (13.5%) had no evidence of extrauterine disease intra-operatively but were found to have a microscopic omental or peritoneal disease on the final histopathology.

### Surgical cytoreduction status

Optimal cytoreduction was achieved in 67.6% (n=25) of patients who underwent primary cytoreductive surgery. This percentage increased to 87.5% (n=7) in patients who received neoadjuvant chemotherapy before cytoreductive surgery. Cytoreduction was suboptimal in 32.4% (n=12) of the primary cytoreductive surgery group, compared with 12.5% (n=1) of the neoadjuvant chemotherapy group.

In five patients, no apparent extrauterine disease was detected intra-operatively, but microscopic omental/peritoneal disease was reported on final pathology. Excluding these patients from the analysis, the rate of optimal cytoreduction among those who underwent primary cytoreduction surgery drops to 62.5% (n=20) of patients. The rate of optimal cytoreduction for patients who underwent primary surgery for a presumed ovarian cancer was 50% (n=3) of patients in the primary cytoreductive surgery group and 75% (n=3) of patients in the neoadjuvant chemotherapy group.

### Survival outcomes

#### Survival according to Histopathological diagnosis

Median overall survival for women with pure uterine serous carcinoma was 16 months, compared with 32 months for women with mixed histopathology (P-value =0.3), regardless of the treatment offered.

#### Survival according to treatment modality

Median overall survival for the cohort was 20 months (8.7–31.3). Patients who underwent primary cytoreductive surgery had an overall survival of 27 months, compared with 20 months for those who underwent neoadjuvant chemotherapy and only two months for those who received only palliative treatments (Table2, Figure1). Progression-free survival was five months for the primary cytoreductive surgery group and six months for the neoadjuvant chemotherapy group.

**Table 2:** Stage IV Uterine serous carcinoma overall survival outcome based on treatment.

| Groups | Subgroups | Mean Overall survival<br>in months | ( 95% CI ) | Median Overall survival<br>in months | ( 95% CI ) |
| --- | --- | --- | --- | --- | --- |
| **PCR ( N=37)** |  | 40.1 | ( 27.3 -52.9 ) | 27 | ( 12.7-41.3) |
|  | Optimal ( N =25) | 43.9 | ( 29.2 -58.6 ) | 36 | ( 13.2-58.8) |
|  | Suboptimal (N= 12) | 27 | ( 8.6-46.6 ) | 15 | ( 0.0 -37.1 ) |
|  | Chemotherapy (N= 33) | 49.4 | ( 34.9 - 64.0 ) | 36 | ( 22.1-49.9) |
|  | No adjuvant CTX (N= 34) | 6.5 | ( 2.4 – 10.6 ) | 4 | ( 2.7 – 5.3 ) |
| **NACT ( N=9)** | NACT ( N=9) | 21.1 | ( 13.5-28.7 ) | 20 | ( 11.2-28.8 ) |
| **PALL ( N=4 )** | PALL ( N=4 ) | 5.5 | (0.2-10.8 ) | 2 | ( 0.0 -6.9 ) |
| **TOTAL** | **TOTAL ( N= 50)** | 33.6 | ( 23.7- 43.5 ) | 20 | ( 8.7- 31.3 ) |

Abbreviation: PCR, Primary cytoreductive surgery; NACT, Neoadjuvant chemotherapy; PALL, Palliative care only; CI, confidence interval

**Figure 1:**
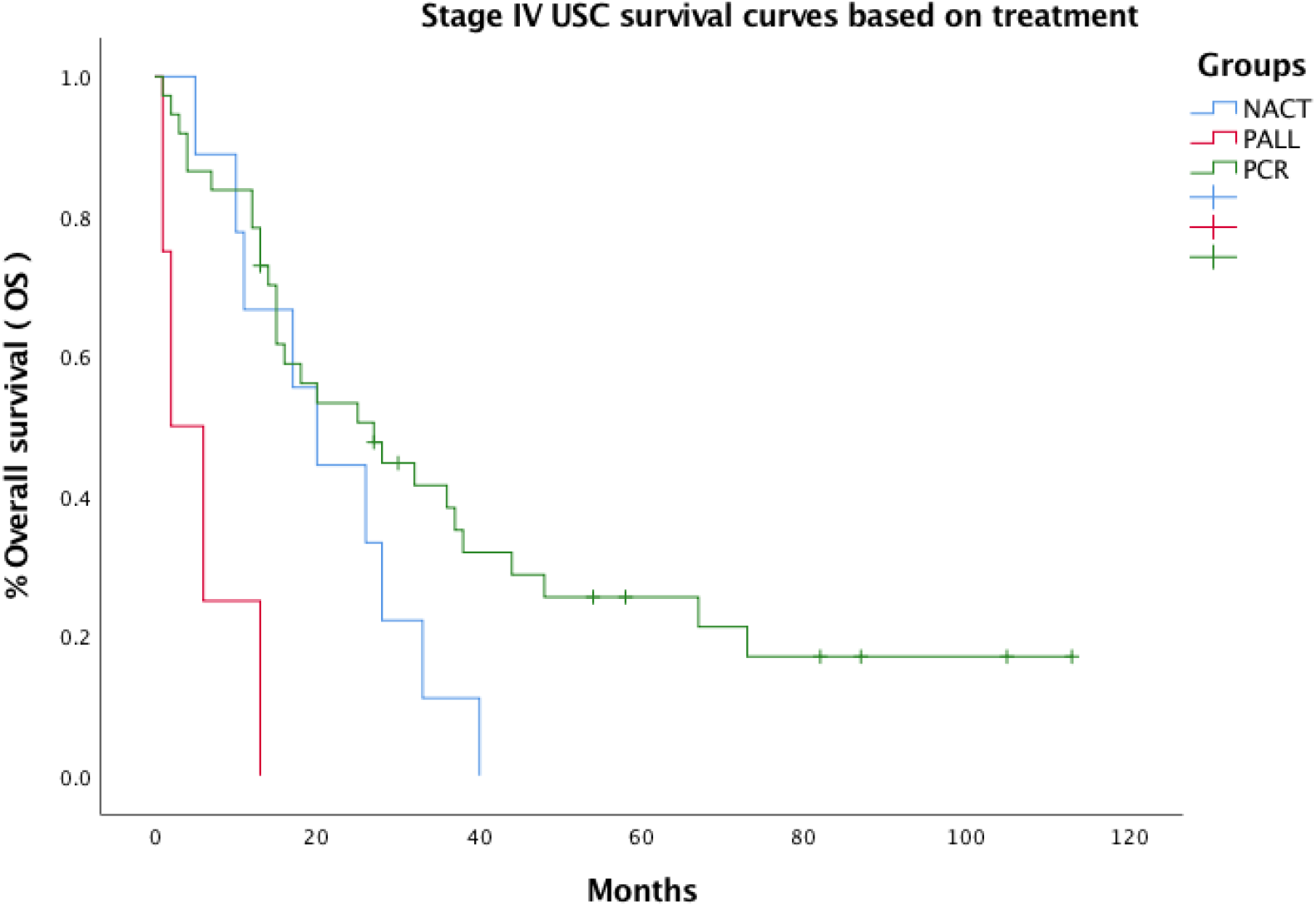
Stage IV Uterine serous carcinoma overall survival curves based on treatment offered. Kaplan-Meir survival curve. Abbreviation: PCR, Primary cytoreductive surgery; NACT, Neoadjuvant chemotherapy; PALL, Palliative care only; OS, overall survival.

Overall survival was significantly longer in patients who received adjuvant chemotherapy after primary cytoreductive surgery, compared to those who did not (36 months versus 4 months, P value <0.05), (Table2, Figure3).

**Figure 2:**
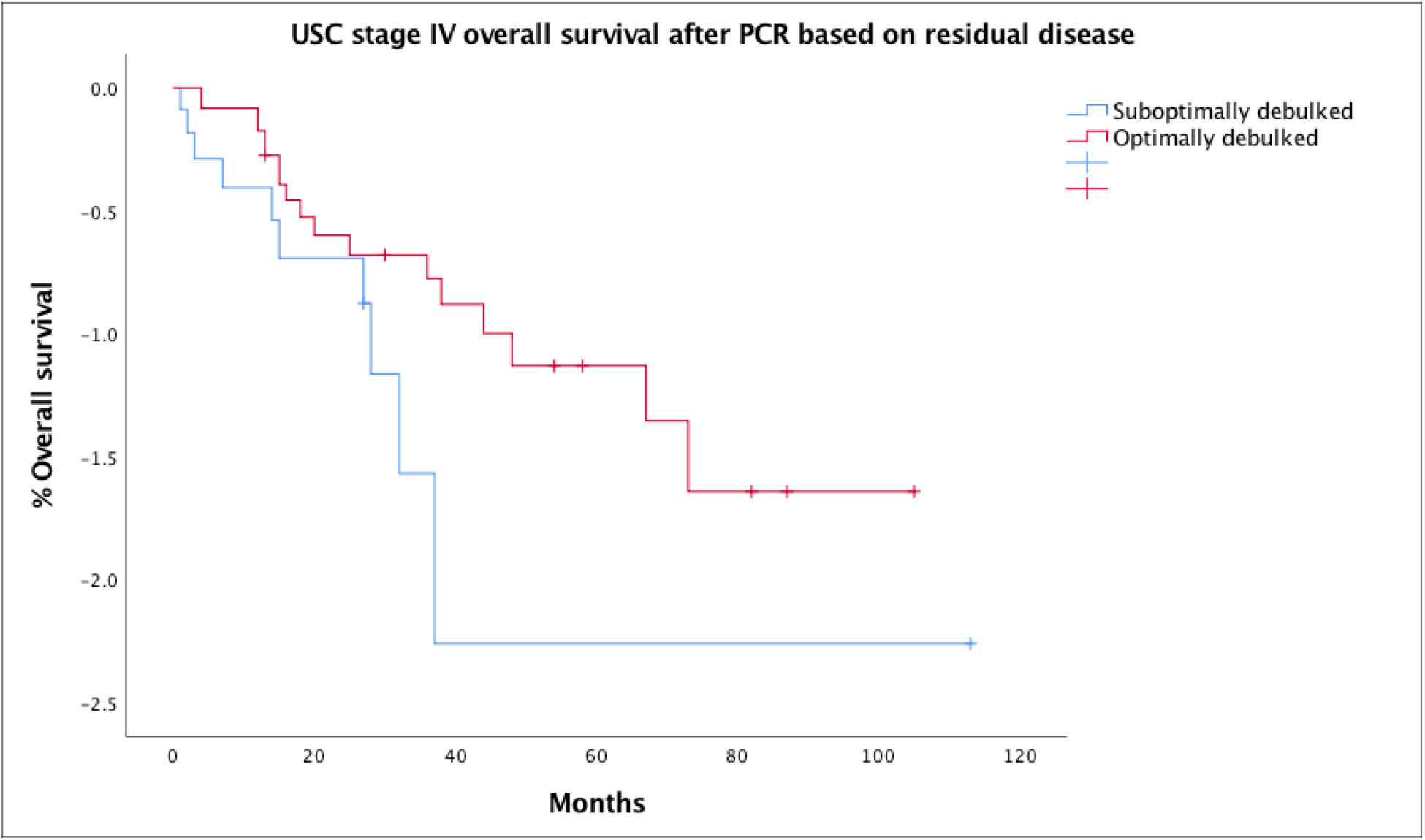
Stage IV Uterine serous carcinoma overall survival outcome based on Cytoreduction status after primary cytoreductive surgery. Kaplan-Meir survival curve. Abbreviation: PCR, Primary cytoreductive surgery, USC, Uterine serous carcinoma

**Figure 3:**
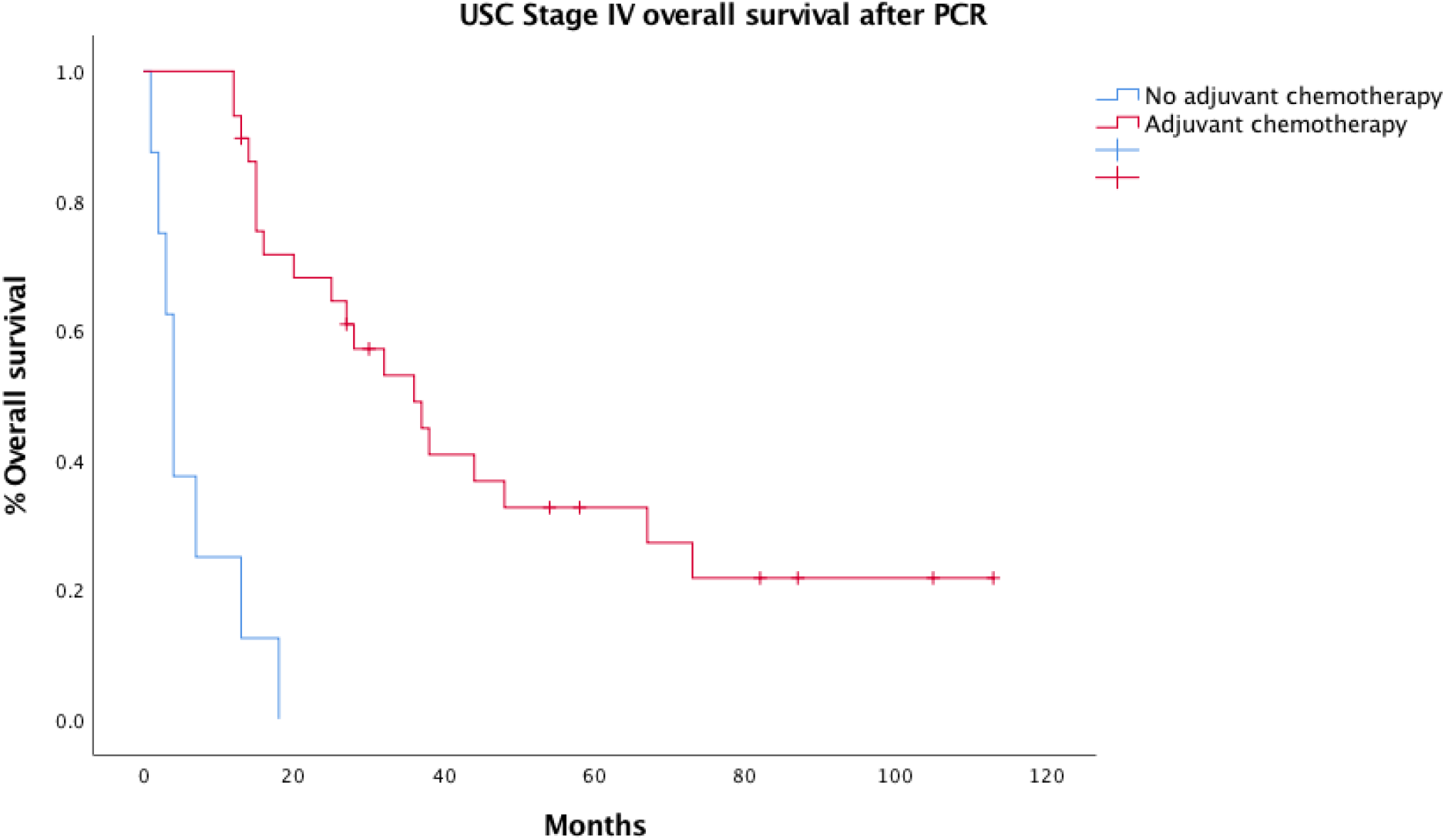
Stage IV Uterine serous carcinoma overall survival outcome based on the use of adjuvant chemotherapy after primary cytoreduction. Kaplan-Meir survival curve. Abbreviation: PCR, Primary cytoreductive surgery, USC, Uterine serous carcinoma

#### Survival outcomes according to cytoreductive surgery status

Among patients who underwent primary cytoreductive surgery, median overall survival was longer in those in whom optimal cytoreduction was achieved than those with suboptimal cytoreduction (36 months versus 15 months, P=0.13), (Table2, Figure2). The overall survival for those who had only microscopic disease was 20 months.

#### Surgical outcomes following Neoadjuvant chemotherapy

Among patients who received neoadjuvant chemotherapy, eight (88.9%) underwent interval cytoreductive surgery. Five patients (55.6%) were found to have a small-volume disease, suggesting an excellent response to chemotherapy. Of the three patients (33.3%) found to have a significant nodular disease, cytoreduction was optimal in two and suboptimal in the other. One patient (11.1%) progressed during neoadjuvant chemotherapy and was not offered surgery.

#### Survival according to treatment era

Of 28 patients treated before the year 2010, two (7.1%) received neoadjuvant chemotherapy, compared with 7 of 22 patients (31.8%) treated between 2010 and 2015. Overall survival was similar for both groups and the overall cohort (20 months).

### Surgical complications

Reported surgical complications were infrequent in the studied cohort. There were no significant postoperative adverse outcomes reported among patients who underwent neoadjuvant chemotherapy followed by interval cytoreductive surgery. There was one postoperative death within 28 days of surgery in a patient in whom the extensive disease was revealed immediately following incision and in whom cytoreduction was not attempted. Other reported adverse outcomes of primary cytoreductive surgery included pelvic hematoma (n=1), atrial fibrillation (n=1), rectal injury/bowel injury (n=2), lymphocyst (n=1), incisional hernia (n=10) and estimated blood loss >1000 (n=1).

## DISCUSSION

Few data are available in the literature to inform the optimal strategy for management of stage IV uterine serous carcinoma, and future randomised controlled trials are unlikely, considering the rarity of the disease and the associated difficulties of recruitment.

Advanced uterine serous carcinoma has biological and clinical similarity to advanced epithelial ovarian malignancy, and may mimic it on initial presentation. In our series, 16.2% (n=6) of the patients underwent primary cytoreductive surgery based on a preoperative impression of ovarian epithelial malignancy, and 44.4% (n=4) of patients who received neoadjuvant chemotherapy were found to have stage IV uterine serous carcinoma on the final histopathology despite the pretreatment diagnosis an advanced epithelial ovarian malignancy. Interestingly, 29.7% (n=11) in the primary cytoreductive surgery group were not suspected to have advanced uterine serous carcinoma disease based on preoperative imaging and were either found to have small volume miliary disease or microscopic peritoneal or omental disease on final histopathology. These findings could be explained by the inherent ability of uterine serous carcinoma to spread early in comparison with other types of uterine cancers. It was interesting to know that even those with microscopic disease upon presentation had a poor prognosis similar to those with gross disease. Mixed histology is very common in type II endometrial cancer^26,27^. Overall survival was worse in pure serous histology, compared with mixed histology.

Surgery remains the mainstay of treatment of uterine serous carcinoma, in the form of total abdominal hysterectomy-bilateral salpingo-oophorectomy and tumour cytoreduction. The surgical approach to primary cytoreduction in advanced uterine serous carcinoma is identical to the approach used in advanced ovarian cancer. In our cohort survival was almost doubled for women who underwent surgery and achieved an optimal cytoreduction, compared with women who did not, although this difference was not statistically significant. The strongest predictor of survival was shown to be the amount of residual disease, although the quality of evidence is lower compared to ovarian cancer research.^4,9,10,14,28^ Failure to achieve optimal cytoreduction appears to carry a worse prognosis for survival in women with uterine serous carcinoma than in those with ovarian cancer.^9^

Adjuvant chemotherapy has been shown to improve survival in several published studies.^12-14,16,29,30^ In our series, all chemotherapy regimens contained at least one platinum component, mostly carboplatin. The survival advantage was 32 months, (36 versus four months; P<0.05), which was the only difference between treatment groups that reached statistical significance.

The value of neoadjuvant chemotherapy has been investigated extensively in ovarian cancer research, where it has been shown to be non-inferior to primary cytoreduction surgery and have the advantage of less perioperative complications.^31-33^ It has been proposed to be the preferred treatment for women with advanced ovarian cancer for whom surgery is unsuitable due to poor performance status or extensive disease with less likelihood of achieving optimal cytoreduction^34 35^. In consideration of the biological and clinical similarities between ovarian cancer and uterine serous carcinoma, Neoadjuvant chemotherapy has been used for patients with stage IV uterine serous carcinoma, with most of the evidence extrapolated from the ovarian cancer research. Reported results in the treatment of uterine serous carcinoma are similar to those of ovarian cancer series, but with lower progression-free survival and overall survival rates.^21-24^

In our series, overall survival was lower in the neoadjuvant chemotherapy group than the primary surgery group (20 months versus 27 months). This difference was statistically nonsignificant and is likely to be due to differences in demographics and patient selection, rather than treatment-related effects alone. The median age and performance status were comparable between neoadjuvant chemotherapy and primary surgery groups, but the neoadjuvant chemotherapy patients had higher comorbidity index compared to the latter. This suggests that patients with high Charlson morbidity index scores were less likely to have been offered or to have accepted surgical treatment. Although the rate of optimal cytoreduction was higher among patients who underwent neoadjuvant chemotherapy who underwent cytoreductive surgery compared to patients who underwent primary surgery, this did not translate to better survival for the reasons above. However, optimal cytoreduction itself was associated with a statistically significant survival advantage in both groups.

No major postoperative complications were reported after interval cytoreduction in the neoadjuvant chemotherapy group, with few reported in the primary surgery group. The small cohort size and the low incidence of complications preclude any comparative statistical analysis of perioperative outcomes.

The patients who received palliative treatments only were older and had a worse performance status and medical morbidity index and had the worst survival among the groups, which reflect the aggressiveness of the disease and its rapid progression without treatment.

In our cohort, Neoadjuvant chemotherapy followed by interval cytoreduction and adjuvant chemotherapy was a safe option for patients with advanced uterine serous carcinoma, achieving a higher rate of optimal cytoreduction and similar overall survival, compared with primary cytoreduction surgery group. Adjuvant chemotherapy was associated with improved overall survival. Pure uterine serous carcinoma was associated with a worse outcome than mixed-type uterine cancer.

Limitations of this study include potential bias associated with retrospective observational data. Some confounding factors could not be captured. The allocation to treatment was physicians depended and based on personal preferences rather than strong evidence. These included the duration of surgery, intraoperative blood loss and length of stay, which were not documented consistently among the group. Although all cases were treated within the Queensland Centre for Gynecological Cancer, operations were performed in nine different hospitals with different medical records systems, which limited the value of comparative analysis.

## Data Availability

The data is confidential but could be accessed by third party if a reasonable request is submitted to Queensland Centre for Gyanecological Cancers

